# A scaling approach to estimate the COVID-19 infection fatality ratio from incomplete data

**DOI:** 10.1101/2020.06.05.20123646

**Authors:** Beatriz Seoane

## Abstract

SARS-CoV-2 has disrupted the life of billions of people around the world since the first outbreak was officially declared in China at the beginning of 2020. Yet, important questions such as how deadly it is or its degree of spread within different countries remain unanswered. In this work, we exploit the ‘universal’ growth of the mortality rate with age observed in different countries since the beginning of their respective outbreaks, combined with the results of the antibody prevalence tests in the population of Spain, to unveil both unknowns. We validate these results with an analogous antibody rate survey in the canton of Geneva, Switzerland. We also argue that the official number of deaths over 70 years old is importantly underestimated in most of the countries, and we use the comparison between the official records with the number of deaths mentioning COVID-19 in the death certificates to quantify by how much. Using this information, we estimate the fatality infection ratio (IFR) for the different age segments and the fraction of the population infected in different countries assuming a uniform exposure to the virus in all age segments. We also give estimations for the non-uniform IFR using the sero-epidemiological results of Spain, showing a very similar growth of the fatality ratio with age. Only for Spain, we estimate the probability (if infected) of being identified as a case, being hospitalized or admitted in the intensive care units as function of age. In general, we observe a nearly exponential growth of the fatality ratio with age, which anticipates large differences in total IFR in countries with different demographic distributions, with numbers that range from 1.82% in Italy, to 0.62% in China or even 0.14% in middle Africa.

## 1 Introduction

The severe acute respiratory syndrome coronavirus 2 (SARS-CoV-2) has quickly spread around the world since its first notice in December of 2019. The pandemic of the disease caused by this virus, the coronavirus disease 2019 (COVID-19), at the moment of this writing, has claimed more than 380 thousand lives. Many countries in the world have declared different levels of population confinement measures to try to minimize the number of new infections and to prevent the collapse of their respective health systems. As the first wave of the outbreak starts to be controlled, the question of how to proceed next arises. The daily number of deaths is progressively decreasing in Europe, and with it, the majority of the countries are starting to release the national lock-downs. The design of future strategies will be sustained on the evolution of the official statistics, and the problem is that these statistics are very defective and incomplete. This is so because, on the one hand, the total number official cases is strongly limited by each country’s screening capacity, which means that only a small fraction of the total infections is correctly identified (typically those presenting symptoms above a certain level of severity fixed by each country’s policy). On the other hand, the shortage of screening tests and an overwhelmed health system also tend to underestimate the number of deaths in the official records. The actual degree of under-counting for both measures is unknown and most likely country dependent, which results in largely irreconcilable case fatality ratios all over the world.

Efforts have been made to determine the clinical severity of the virus [1, 2, 3], but determining precisely how deadly this virus is remains hard [4]. Many different solutions using the available data have been proposed to extract the correct case fatality ratio [5, 6], estimate the number of infections [7, 8] or the infection fatality ratio [9, 10, 11, 12]. Even the results of some early sero-epideomiological tests sampling the population degree of immunity have been strongly controversial [13, 14]. Probably the most reliable estimations for the infection fatality ratio (IFR, the probability of dying once infected) as a function of the patient’s age, were proposed by Verity *et al*. in Ref. [10] using the data from 4999 individual cases in mainland China and exported cases outside China. The ratios obtained were further validated with the reported cases in the Diamond Princess cruiser. Yet, these estimations were based on two assumptions. Firstly, a perfect detection of all the infections among people in their fifties, a debatable hypothesis given how elusive the detection of this virus is. And second, that the virus had spread uniformly within the population of all ages, which is rather improbable in their case because they were analyzing mainly infections among travelers (that tend to be younger). Nevertheless, the picture is clear, the lethality of the virus increases sharply with the patients’ age, being particularly deadly for elderly people and mild for kids.

In the absence of a reliable number of confirmed infections, most of the statistics have focused on the number of deaths, which are expected to be a fraction of the first one. But deaths are much less common than infections, which means that in order to estimate correctly the number of infections of a country, one needs a very accurate death counting. In this sense, it is widely accepted that the number of real deaths linked to COVID-19 is noticeably larger than what officials statistics say [15, 16], but estimating precisely how much is hard and will likely depend strongly on the country data collection policy and capacity. One can try to estimate the size of this discrepancy from the excess mortality observed since the beginning of the pandemic in the public death records. This approach, though apparently infallible, is not without difficulties. Indeed, in most of the countries, the epidemic peak took place at the same time as that of lock-down measures, which means that, on the one hand, the mortality for other reasons (not COVID-19) has decreased, and on the other hand, the health system being under a lot of stress, the mortality linked to other diseases has increased. Correcting these effects in the reference mortality trend requires a careful an exclusive analysis. We reason in an alternative way.

The under-counting of deaths comes from mainly two sources: (i) only the deaths that can be directly linked to COVID-19 (by means of a positive result in a PCR test, typically) are included in the official counting and (ii) countries mostly count the deaths occurred within hospital facilities in the statistics. Source (i) tells us that all the patients that die before being tested are invisible. This will happen eventually at all ages but since old patients are more prone to develop severe symptoms and have more difficulties to seek immediate medical attention, this situation will be far more common among the elderly. Also source (ii) mainly affects old people because being hospitals crowded, the oldest patients have been often treated in retirement/care homes or in their own homes. For these reasons, we expect a significantly more accurate reporting of the deaths of younger patients (in particular, under 70 years old). It is possible to quantify this idea.

According to the Office of National Statistics in the United Kingdom, among deaths mentioning COVID-19 in the death certificate (in England and Wales by the 22nd of May) 64% took place in hospital, 29% in care houses and 5% at home [17]. Analogous data published by the Community of Madrid’s government (which counts more than 1/3 of the official deaths in Spain) reports similar ratios: 61% hospitals, 32% socio-sanitary places and 6% home. France counts separately the deaths occurring in hospitals and in care homes, and the latter being almost 60% of the former. Deaths occurring in care houses are a large portion of the total in all countries, which means that an incomplete counting there, modifies notably the overall statistics. However, once we look at the mortality per age group, such under-counting only affects the patients of a certain age. In fact, we can compare the number of deaths having COVID-19 mentioned in the death certificate (even if it is only a suspicion, which most probably represents an over-counting of the real deaths) and the official counting of deaths linked to COVID-19. In Fig. 1, we show the excess of the former with respect to the latter, relatively to it (that is, suspected deaths divided by the official deaths minus 1) for England and Wales and the Community of Madrid. In both places, the under-counting is relatively age independent under 70–80 years old, and rather important above, specially for the patients above 90 years old, where real numbers may probably double the official counting. Furthermore, this mismatch is getting worse as records in England and Wales are correctly updated (in Madrid it seems rather stabilized). Details on the data used to generate these plots are given in the Methods and Dataset section.

**Figure 1:**
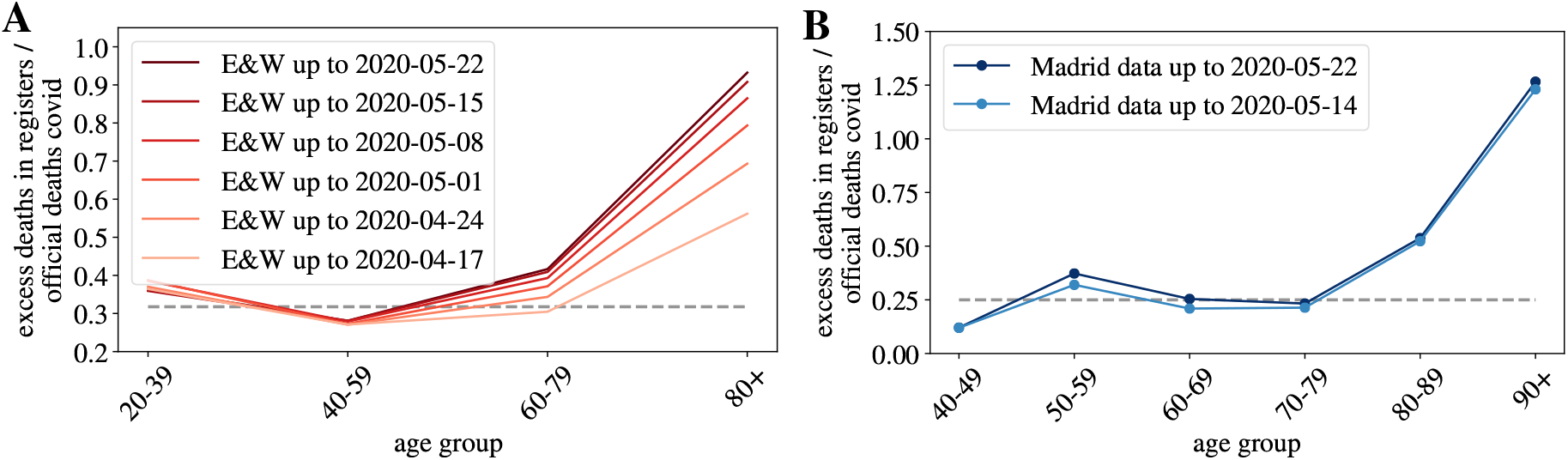
Under-counting of deaths per age groups.. We show the excess deaths, per age groups, observed when comparing the number of deaths certificates where COVID-19 was mentioned either confirmed or suspected, and the official deaths attributed to COVID-19, relatively to this second number, for England and Wales in A, and for the Community of Madrid B.

In summary, we expect a small mismatch between the real and the official number of deaths among patients under 70 years old (the *∼* 30% of under-counting is probably too large because deaths caused by other diseases are probably also included in this count), and a much higher systematic under-counting for the older segments. The actual numbers will depend on the country capacity to detect quickly the infections, but also on the particular details concerning the counting of official deaths (which establishments are considered). We give these details, together with the last date used for each country in the Methods and Dataset section.

In this work, we attempt to estimate the IFR as function of age using scaling arguments relating the accumulated number of deaths reported in different countries and age groups. We define all our variables in Section 2. We establish a direct correspondence between the mortality rates in patients below 70 years old (where the official counting is more accurate) published in different countries around the world (but mostly in Europe) in Section 3.1. This good correspondence allows us to make predictions about the degree of spread of the virus in different populations, or the global IFR of a country, as compared to another one. We also observe that the collapse of the mortality rate with age in different countries is compatible with a pure exponential growth of the IFR with age (assuming a uniform attack rate). The scale of total infections is then consistently fixed from the rate of immunity obtained via blood tests of a statistical sampling of the citizens Spain in Section 3.2 (and compared to seroprevalence tests in Geneva, Switzerland, and New York City, United States). This scale allows us to compute the IFR as function of age and the number of current infections in each country that are given in Table 2. In addition, we estimate the probability of being detected as official case, needing hospitalization and intensive care (if infected) as function of age in Spain in Section 3.3. All these rates are obtained under the assumption of a uniform attack rate, an assumption that seems fairly reasonable seeing the immunity measures of the Spanish test, measures that, when once taken into account, do not change qualitatively the results discussed so far (see in Section 4.1). Finally, we estimate the dimension of the under-counting of deaths among the elderly in the different countries and give estimations for the overall lethality of the virus in Section 4.2. We relegate all the details concerning the databases and dates used in the data-analysis for the Section 6.

## 2 Definitions

Statistical offices and health institutions of many countries have been publishing regularly the age distribution of the accumulated number of deaths occurred in their territory since the beginning of the outbreak. We have combined national data from Denmark, England& Wales, France, Germany, Italy, South Korea, Netherlands, Norway, Portugal and Spain, regional data from Geneva (Switzerland) and Madrid (Spain), and city data from New York City (Unite States of America). Unless something else is mentioned, we will consider 10 age groups, each gathering together the deaths of patients with ages in the same decade (with the exception of patients over 90 years old are grouped together). Since the different age segments are not uniformly populated, and this distribution can change significantly from one country to another, we will discuss always the number of deaths normalized by the number density of people *x_α_*(*𝒞*) in each age group *α* and country *𝒞*, that is,

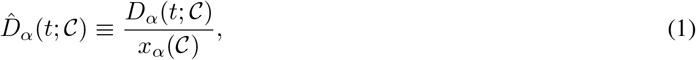

being *D_α_*(*t;C*) the accumulated number of deaths at a time *t*. This variable 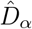 normalized by the country’s population and multiplied by 1000 (by convention) is the *mortality rate* per age group for the time elapsed since the beginning of the outbreak. In the following we omit the country variable *C*, unless explicitly needed. We show in Fig. 2–A the evolution of 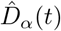 in France for our ten age groups. As shown, once the effects of the demographic pyramid are removed (the fact that there are much more people in their fifties than in the nineties in any population, for example), the mortality expands over almost five orders of magnitude between kids and elderly people.

**Figure 2:**
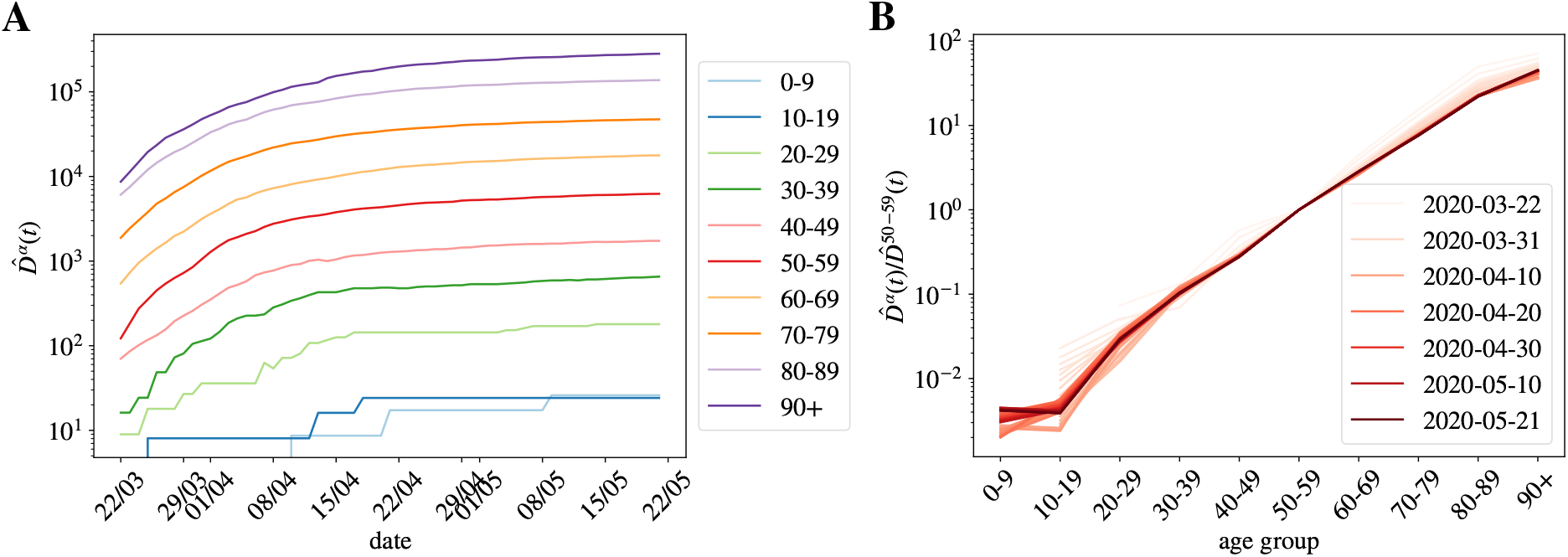
Normalized number of deaths occurred in French hospitals as a function of age. A We show the evolution with time of the accumulated number of deaths normalized by the number density of individuals in age group *α* (i.e. 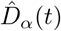 in Eq. (1)). In B, we show 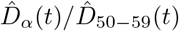 as function of the age group, for all the times in A (the darker the color, the more recent the measurement, and we give some dates in the legend). This quotient is essentially time-independent as discussed in Eq. (6), and it lets us estimate the quotient between the UIFR of the two age groups, that is,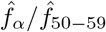

Asymptotically, the accumulated number of deaths in each *α* at a given *t*, will be a fixed fraction of the accumulated number of infected individuals in that group, *I_α_*, at a previous date *t −* Δ, thus Δ is an effective time related to the time elapsed between infection and death (estimated to be, in average, around 20 days [18, 19])^1^. Then,

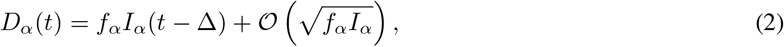

being the proportionality factor, *f_α_*, the infection fatality ratio (IFR) for the age group. The fluctuations *O* are the expected error of an un-normalized histogram. The assignation of a unique delay for all the cases is, of course, an over simplification, but which yet works quite well as the number of infections becomes large. We show, for instance, the perfect match in time between the accumulated number of cases and deaths at a later time in Spain in Fig. S1.

In general, we do not know either the total number of infections *I* =Σ_*α*_ *I_α_*, or the number of infections in a particular age segment *I_α_*, but we know the latter should be an (essentially constant) fraction of the total number of infections, plus fluctuations, that is,

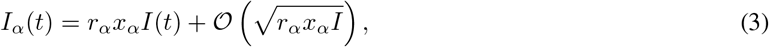

with *r_α_* being the relative risk of infection for group *α* (thus being *r_α_I/N* the standard attack rate for the group, with *N* the total country population). Clearly,Σ_*α*_ *r_α_x_α_* = 1. Recent results analyzing the spread of the virus within close contacts in the outbreak in China suggest a uniform exposure across the population [20], meaning that *r_α_* = 1 for all the groups (quite different from the patterns observed for the seasonal flu [21, 22]). There is, however, an important debate whether the low fatality observed in patients below 20 years old is related to a low risk of death or a low risk of infection. For the moment we keep this variable free and we will discuss it at the end of the paper. The risk of infection *r_α_* could, in principle, vary with time, but we do not observe a systematic change with time. This will be clearer with the discussion around Fig. 2–B for the accumulated deaths, or for the analogous figure concerning the daily measures (which should be more sensitive to a change in *r_α_*) in Supplemental Fig. S2.

## 3 Results

### 3.1 Scaling between age segments

The combination of Eqs. (2) and(3) tells us that:

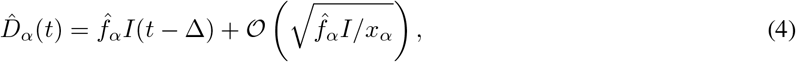

where

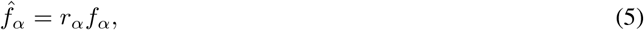

would be the probability of dying with age *α* if the virus attacked uniformly all ages within the population. In other words, it is the “apparent” fatality (what we perceive from the daily news) without knowing if all ages have the same chances of getting infected. For this reason, we refer to 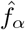 as the *uniform* infection fatality rate (UIFR), as compared to *f_α_*, which is the *real* (potentially non-uniform) IFR associated to the disease. Both measures are only equal if *r_α_* = 1 for all *α*.

All together, for all age segments, 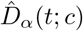 is expected to be proportional to the total number of infections at a previous date, *I*(*t −* Δ). Alternatively, the quotient between the mortality rate of two distinct age groups,

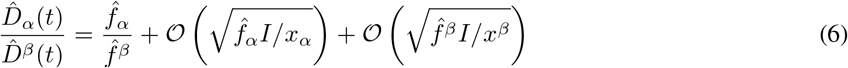

is expected to become independent of time (as long as the number of the expected deaths for each group is large enough), and equal to the quotient between the UIFR of each group. This is precisely what we observe for the deaths occurred in French hospitals (see Fig. 2–B) where we show the quotient between each 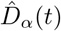, and the deaths among patients in their fifties, 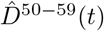 for all daily reports since the 22nd of March of 2020 (the darker the color the more recent the measurements). The other countries considered shows qualitatively the same behavior, we decided to show France because it has been reporting age statistics (on a daily basis) for the entire number of deaths occurred up to that date. Thus, with this kind of analysis, even if we do not know the exact mortality associated to the virus, we can determine how deadlier it is, at least apparently, for an age group as compared to another. We say apparent, because up to here, we cannot distinguish if the virus seems less aggressive for an age segment because the lethality is low or because so few individuals of that age got infected.

The same kind of arguments apply to data from different countries at a fixed time. Indeed, one expects that the IFR, *f_α_*, should not vary too much from country to country (at least within countries with comparable health systems). However, the relative attack risk *r_α_* may vary from country to country. Yet, if the differences in *r_α_* are not large between populations, then, also 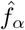 should be country independent. In such case, Eq. (4) tells us that the different 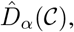, essentially differ by a multiplicative constant proportional to the total number of infections *I*(*𝒞*) in each country. We show in Fig. 3–A, the counting 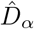 by the 22nd of May of 2020 available for the different countries where we found information about the profile of deaths by decades of age (see Database section for details) as a function of *α*. Some countries publish this data only for a fraction of all the deaths, if this is the case we renormalized the numbers with total number of deaths reported by each country by the 22nd of May of 2020.

**Figure 3:**
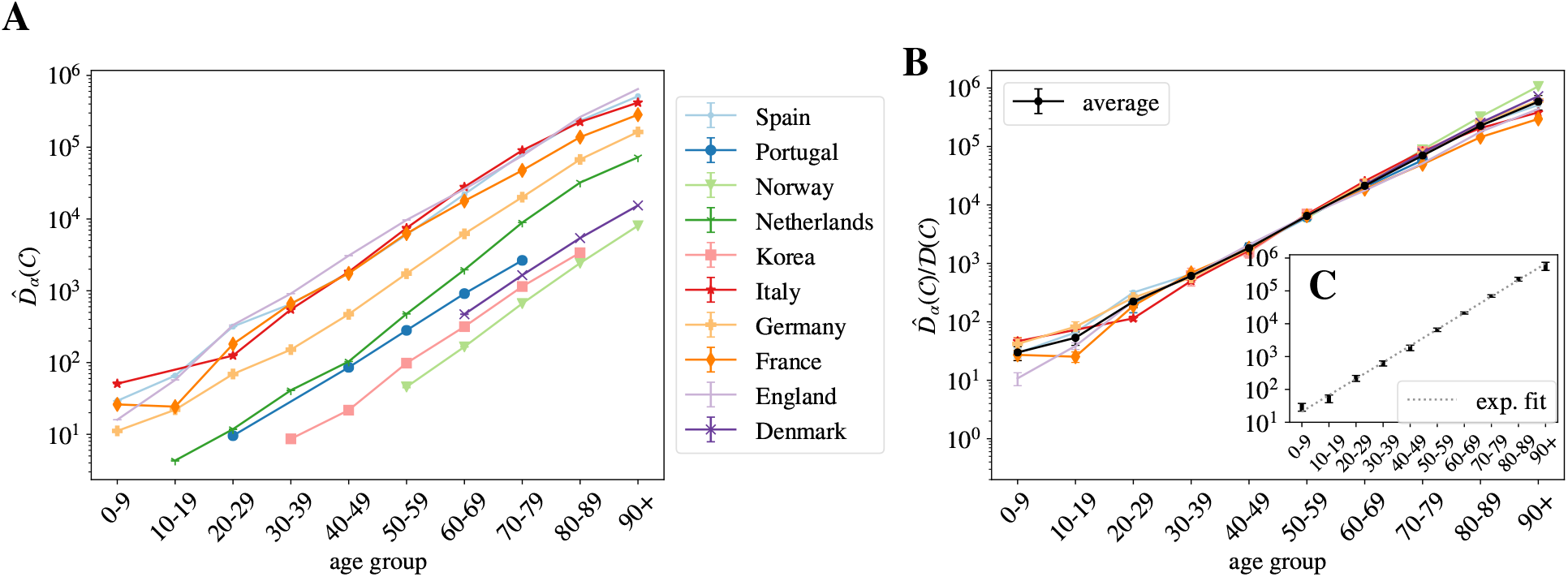
Normalized number of deaths in different countries as a function of age. A We show the normalized number of deaths per age group for a selection of countries affected by the COVID-19 epidemic in very different degrees. In B same data (excluding Netherlands) but where every country has been multiplied by a constant *𝒟*(*𝒞*) so that it collapses with the Spanish curve in the age region in between 30–69 years old. The values of these constants are given in Table 1. In black, we show the country average for each age segment, and in C the fit of this average to a pure exponential.

As argued, the different countries’ curves are essentially parallel in logarithmic scale, with the exception of the Netherlands, where the mortality grows even more steeply with age than in the rest of countries (maybe related to a significantly different *r_α_*). In other words, we can extract both the number of total infections and the UIFR by age (but for a multiplicative constant common to all the countries, or all the ages, respectively) from the collapse of these curves. We show in Fig. 3–B this collapse (where Netherlands was excluded), which works extremely well for all the countries in the age region between 30–69 years old (despite the different orders of magnitude of 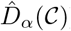. Deaths below 30 are very rare, so strong fluctuations between countries are expected. The collapse is less satisfying above 70 years old, but, as discussed, we believe this is mostly related to a different degree of under-counting of deaths for these segments of age (though differences related to a relative under-representation of elderly people among the infected in some countries are also possible). We believe it is mostly under-reporting because, for instance, the French curve would quickly match the rest of the countries if one added (for the segment over 80 years old) the official deaths occurring in care houses to the hospital deaths shown here. We will try to estimate the extent of this under-reporting in each country below.

One can now exploit this similarity in the growth of mortality with age between countries to remove the statistical fluctuations. Thus, the country average of this collapse gives us the UIFR (but for an unknown proportionality constant 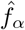 common to all age segments). We give the values of this average in Table 1 (errors are obtained using the boostrap method at 95% of confidence). Data obtained is compatible with an exponential growth of the mortality rate with age (as shown in Fig. 3–C. In fact, a fit of the data to

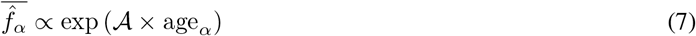

with *A* = 0.115(7) has a *χ*^2^*/*d.o.f = 3.8*/*8. This strong dependence of the fatality with age anticipates that one should expect that the global UIFR 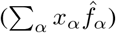 varies a lot from one country to another due to the different demographic distributions. We will discuss this below.

**Table 1:** Collapse of the mortality rate in different countries. We give the values extracted from the collapse of Fig. 3–B: the growth of the mortality with age (proportional to the uniform fatality ratio 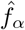) and the number of infections in each country with respect to the number of infections in Spain *I_𝒞_/I*_Spain_ equal to the collapsing constant *𝒟*(*𝒞*). The relative scaling of the mortality above 70 years old is expected to be significantly underestimated. The errors of *𝒟*(*𝒞*) are only the statistical errors extracted from the data collapse, they do not include the systematic error associated to the different policies of death counting the different countries which would be much larger, we try to give a better estimate below.

| Scaling relations |  |  |  |
| --- | --- | --- | --- |
| age group | $\propto \hat{f}_\alpha$ | Country | $D(C) = I_C/I(\text{Spain})$ |
| 0-9 | 28(9) | Spain | 1.0(0) |
| 10-19 | 51(14) | Portugal | 0.045(2) |
| 20-29 | $21(4) \times 10$ | Norway | 0.0076(4) |
| 30-39 | $60(10) \times 10$ | Korea | 0.014(2) |
| 40-49 | $18(3) \times 10^2$ | Italy | 1.1(2) |
| 50-59 | $63(8) \times 10^2$ | Germany | 0.27(2) |
| 60-69 | $21(1.6) \times 10^3$ | France | 0.96(8) |
| 70-79 | $69(7) \times 10^3$ | England | 1.48(2) |
| 80-89 | $22(3) \times 10^4$ | Denmark | 0.02(-) |
| 90+ | $5.6(16) \times 10^5$ | | |

Furthermore, the collapsing constant is essentially the relative number of infected people with respect to our reference country, that is *I*(*C*)*/I*(Spain). This is not entirely true due to the different country policies concerning the death-counting, but, as discussed, we estimate that the unreported fraction under 70 years old is inferior to 30% and the quotient of the underestimation of the two countries would, in general, much smaller. We show these collapsing constants in Table 1.

### 3.2 Fixing the scale

#### 3.2.1 Number of infections and uniform fatality rate

Up to this point, we have only obtained the number of infections by country with respect to the number of total infections in Spain, and something proportional to the UIFR by age. In both cases, the proportionality constants (though both related) are unknown. In order to fix the scale, we can look to the statistical studies of prevalence of antibodies against SARS-Cov2 in different populations. In particular, we refer to the preliminary results of the sero-epidemiological study of the Spanish population (inferred from 60983 participants) made public by the Spanish Health Ministry the 13th of May of 2020 [23], that estimates that only a 5.0% (95% interval of confidence (IC): 4.7%-5.4%) of the Spanish population had been infected (using blood tests drawn in between 27/04–11/05/2020). Also, as an independent control of the scale, we use the results of an analogous seroprevalence survey of the residents of the Geneva, Switzerland (from 1335 participants) [24].

The sampled rate of immunity in the Spanish population allows us to fix directly *I*(Spain) in Table 1 and with it, estimate the number of infections in each of the countries of Fig. 3 (see Table 2). The results obtained are lower, but compatible, with the independent estimations by Phipps et al. [8] or Salje et al. for France [25]. As shown, the rates of infection (for the entire country) are rather low, in particular compared to the 60–70% herd immunity threshold (even if it is lowered for other effects [26]). Yet, it is important to stress that the propagation of the virus has been rather heterogeneous in the territory, being the contagion high in certain regions and insignificant in others. We take for example France, where the age distribution of the COVID-19 deaths is available for all the departments. Using also the data up to the 22nd of May, we estimate that the percentage of the population infected has reached 12% in the Island of France (the department of Paris), 7% in the Great East, 2.5% in Upper France, and it is 1% or less in the rest of departments.

**Table 2:** Estimations assuming a uniform attack rate. We show our estimation for the uniform fatality rate before and after quantifying the effects of the systematic under-counting of deaths. We estimate the percentage of the population infected in each country. Errors include the statistical error (*±sigma*, the standard deviation obtained through error propagation of the results in Table 2, and the uncertainty of the prevalence survey in Spain) and a systematic error of 35% of possible under-counting of deaths).

| age group | Uniform infection fatality rate |  | % Population infected |  |
| --- | --- | --- | --- | --- |
|  | with under-counting | estimation without under-counting | Country |  |
| 0-9 | 0.0012%(4) | 0.00118%(0.00082-0.0016) | Spain | 5.0%(4) |
| 10-19 | 0.0021%(7) | 0.00211%(0.0014-0.0028) | Portugal | 1.0%(4) |
| 20-29 | 0.009%(23) | 0.00878%(0.0065-0.012) | Norway | 0.33%(12) |
| 30-39 | 0.024%(5) | 0.0241%(0.019-0.032) | Korea | 0.06%(2) |
| 40-49 | 0.072%(18) | 0.0722%(0.056-0.097) | Italy | 4.3%(16) |
| 50-59 | 0.26%(5) | 0.256%(0.21-0.35) | Germany | 0.8%(3) |
| 60-69 | 0.84%(0.14) | 0.839%(0.71-1.1) | France | 3.4%(12) |
| 70-79 | 2.8%(5) | 3.47%(2.9-4.7) | England | 6%(2) |
| 80-89 | 8.9%(18) | 12.7%(11.-17.) | Denmark | 0.9%(3) |
| 90+ | 23.%(7) | 42.1%(34.-57.) |  |  |

Furthermore, the total number of infections lets us compute the UIFR as function of the age in Spain just by dividing our 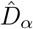 by this number, that is, using Eq. (2),

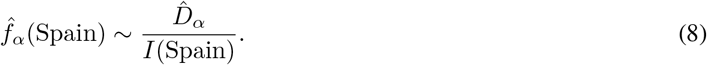

We show the values obtained in Fig. 4–A. Then, we can extract 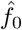 from the comparison of ^*f_α_*(Spain) with the values 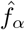 in Table 2, in the age regions where we believe that the counting of deaths is reliable (the region where the collapse of Fig. 3–B was good). We use the group 50–59 to fix this constant 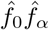, which allows us to reconstruct entirely our estimate for the averaged UIFR (we show these values in Fig. 4–A and Table 2). This determination of the UIFR is expected to underestimate the fatality ratio for the oldest segments of data, we will try to correct this bias in the next section. We also include this second estimation in Table 2).

**Figure 4:**
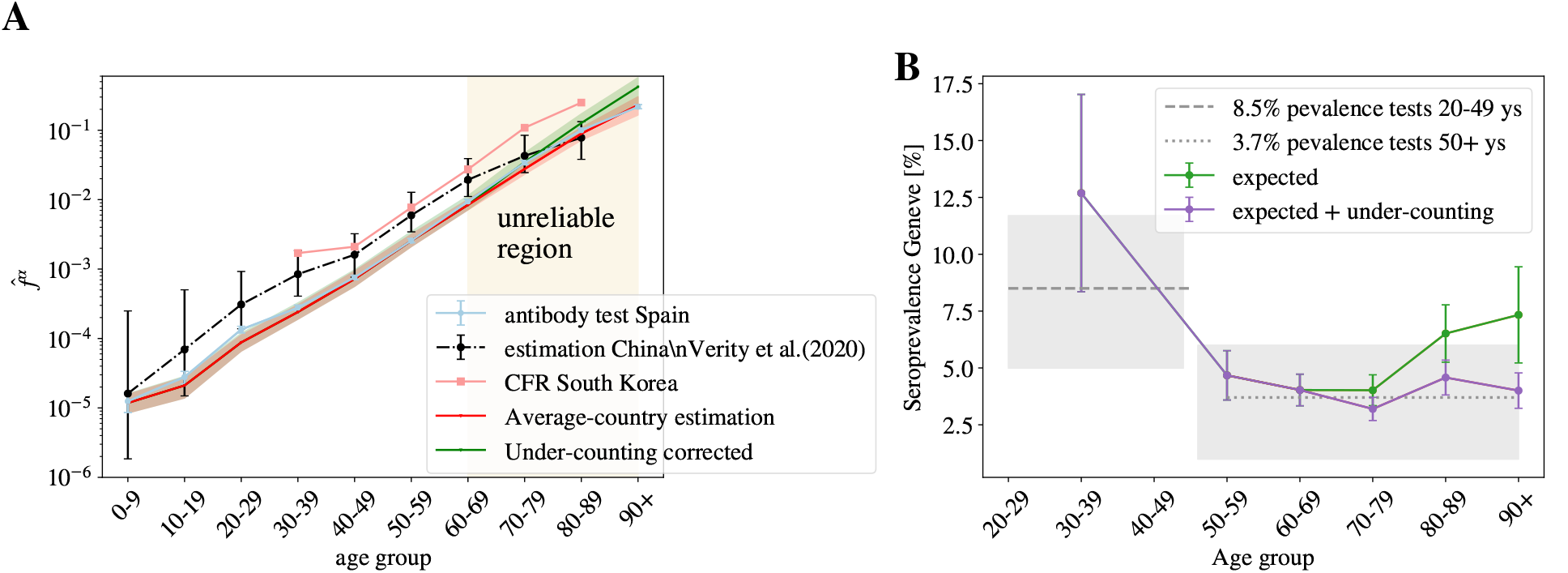
Probabilities assuming a uniform attack rate. A We use the measurements of the number of infected in Spain to estimate the uniform fatality rate using Eq. (2) in both regions. We fix the constant ^*f*_0_ in Table 1 using the estimation of the fatality in Spain for the age group 50–59 to infer the average UIFR. We show this estimation in red and in green we show our estimation of the UIFR after the under-counting of deaths in the old segments have been corrected. We compare these results with the estimation by Verity *et al*. [10] and the case fatality ratio (i.e. the probability of dying for confirmed COVID-19 cases, not the IFR) by age obtained in South Korea. In B, we use the IFR estimations from A and Table 2, to predict the seroprevalence of anti-SARS-CoV-2 antibodies in the population of Geneva, Switzerland, from the official distribution of deaths per age of a total of 277 deceases. The predicted fraction of infections is given in dots (in green, if we used the bare estimation of Eq. (8), in violet, if we include the corrections linked to under-counting). In horizontal lines (and the 95% of confidence interval in gray shadow), we show the actual values measured from the survey of Ref. [24] for patients of different age-groups.

We can test the accuracy of the estimated IFRs by this method using another independent sero-epidemiological survey. In particular, we use the work by Stringhini *et al*. [24] that measures the degree of seroprevalence in the canton of Geneva (Switzerland) from samples of 1335 participants. Up to the 24th of May of 2020, the canton’s authorities had reported 277 deaths, all but one from patients above 50 years old. We can use the age distribution of these deaths and our estimation of the IFR in Table 2, to guess the fraction of the population that have been infected so far using Eq. (2). We show in Fig. 4–B, the quotient 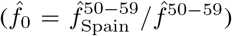, being *N* the total population of the canton of Geneva. If our 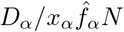 is, indeed a good estimation for the real IFR, this quotient should give us the fraction of the population infected in that age group, which was estimated to be very similar above 50 years old and equal to 3.7% (95% CI 0.99–6.0) and about 8.5% (95%CI 4.99–11.7) in between 20–49 years old [24]. As shown, our predictions are in very good agreement with the survey estimation (specially once the systematic under-counting of deaths in the estimation of the IFR is corrected, see Section 4.2).

The convergence of the results in Spain and Switzerland lends great confidence to the ratio between deaths and infections, but let us stress that these estimations might be only valid for similar health systems, and for hospitals not too overwhelmed during the worst moments of the epidemic peak. In fact, if we use the IFR of Table. 2 to estimate the percentage of infections in New York City from the distribution of the deaths with age published by NYC Health at different dates (we show the results per age in the Supplemental Fig. S3), we obtain predictions for the overall antibody prevalence that evolve in time from 27% (data from 15th of April), 48% (the 1st of May), 57% (the 15th of May), to 63% (the 2nd of June), which would indicate that herd immunity would had already been reached in the city. However, there are proofs that this is not the case. Indeed, the presence of antibodies within the NYC’s citizens was randomly sampled, at a certain point at the end of April (details have not been published), in the base of a survey of 15000 people in all the New York State. The results announced by the Governor in a press conference the 2nd of May of 2020 reported that only a 19.9% of the tested had antibodies. If we move forward *∼* 20 days in time to see this reflected in the deaths [18, 19], we estimate 3 times more infections, which inevitably suggests that the infection fatality rate have been much higher in New York City that what it was in Spain or in Geneva, unless there are issues in the sero-prevalence study, something hard to estimate because technical details of the survey have not been published so far (to our knowledge).

We can also compare our IFR with previous estimations. Our numbers are smaller than the estimation by Verity *et al*. [10] for all the age segments except those that concern the elderly patients (though still compatible with their confidence interval for most of the age groups), and about three times smaller than the case fatality ratio (the probability of dying among the confirmed cases) per age group measured in South Korea (where a massive number of screening tests has been made). This difference could be explained, in both cases, from an under-estimation of the number of total number of infections. On the one hand, the IFR in Ref. [10] was estimated from the case fatality rate, and the statistical prevalence of antibodies among the travelers returning home from repatriation flights (which represents a much lower sampling that the one considered in the Spanish survey). On the other hand, Korea has been very successful identifying new infections by tracking the social contacts of the infected, but it is very unlikely that they are able to trace all the infections.

Before ending this Section, we would to warn about the limitations of the current sero-epidemological surveys, which will probably affect our results (even though we would like to stress that the Spanish survey has been praised for its robustness [27]). In fact, extracting accurate results from them is challenging for different reasons. Firstly, because the study must be well designed to avoid undesirable bias in the recruitment of the participants. Secondly, because the probability of detecting the antibodies change with time [28] (an effect that must be taken into account [29]). Thirdly, because available tests are not very accurate [30], which means that statistical adjustments must be included in the analysis to avoid mistaking the antibody rate with the false positive rate [31]. And finally, because the spread of the virus have been very heterogeneous in space (as we illustrated for France above), which means that very large samples are necessary to get the correct picture of a country.

### 3.3 Other probabilities

Spain also gives age distributed data (for groups of patients with ages in the same decades) for the accumulated number of cases, *C_α_*, new hospitalizations, *H_α_*, and new admissions in intensive care units, *S_α_*. Due to the shortage of screening tests, for most of the age groups, the number of cases gives us a measure of the number of patients with symptoms severe enough to visit an emergency room. For the oldest groups, it might not be the case because care houses with confirmed cases have been more systematically tested than the rest of the population. Then, we apply the same reasoning used to compute the UIFR to these indicators, which allows us to estimate the probability of being included in each of the other three categories. Unlike the deaths, policies concerning who get tested, hospitalized and/or admitted in an intensive care unit probably depend strongly on the country, which means that these probabilities might not be directly extrapolated to other countries.

Equation (4) reads for a general observable *X* (*X* = *C, H, S*, or *D*),

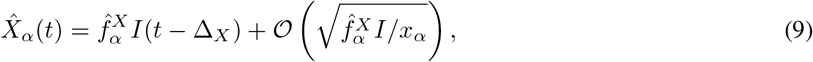

which means that we can directly extract the probability of being included in the *X* category 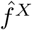(again assuming a uniform attack rate) using the measure *I*(Spain) from the antibody prevalence study [23]. Note that knowing the precise value of Δ_*X*_ is not crucial here because the propagation of the disease has been mostly interrupted in Spain during the last month, and *I*(*t −* Δ_*X*_) is roughly constant at this point. We show the estimations of these probabilities per age group in Fig. 5.

**Figure 5:**
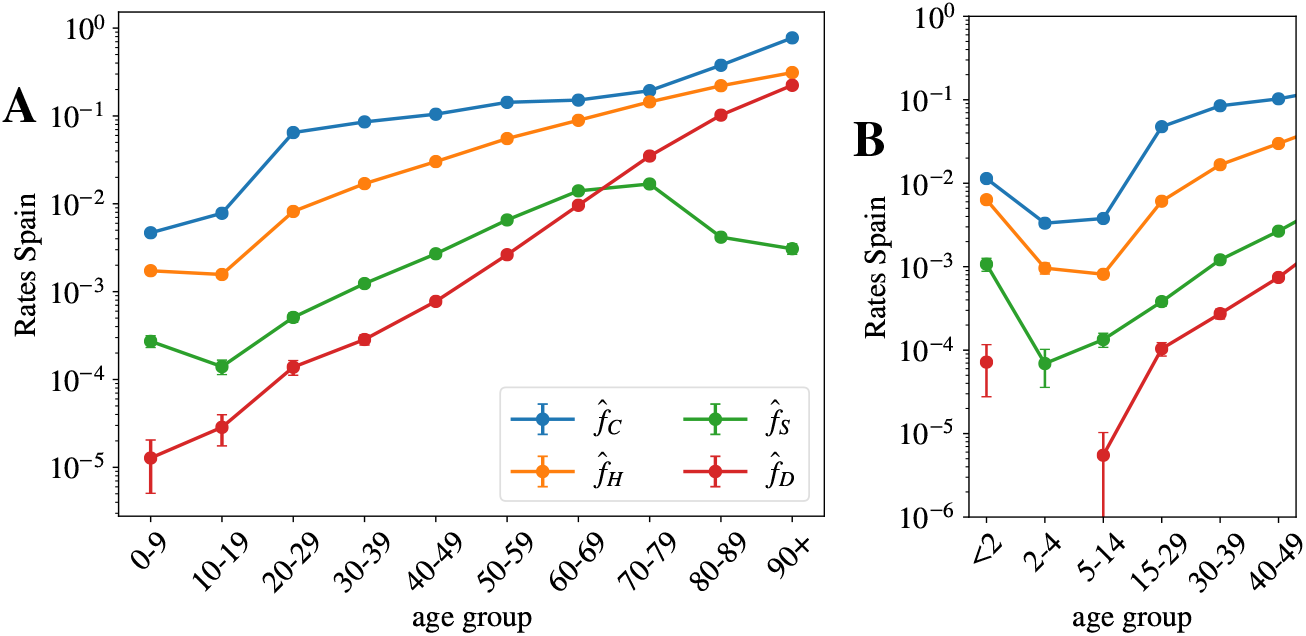
Other probabilities assuming a uniform attack rate. In A we show the probability of being classified as official case,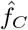 being hospitalized, 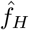 admitted in intensive care, 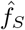, and dying, 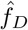, in Spain, as function of the age using age segments of 10 years. B, we show the same data but were the kid’s information has been grouped by smaller age-segments, evidencing the severity of the cases in patients under 2 years old. A is generated using the data by the Spanish Health Ministry up to the 22nd of May and B with the data published by the RENAVE.

We see that, between 20–80 years old, the probability of being confirmed as a case does not depend too much on age, and it keeps fixed around 1 every 10 infections. The probability is higher for older segments and much smaller for people below 20 years old. For the other indicators, we observe a strong dependence of the severity with age. For the intensive care unit admissions, however, above 70 years old, one sees clearly the effects of the policies regulating the access to intensive care with age, an access that becomes rare over 80 years old. A situation which certainly contributes to increasing slightly the mortality rate for the oldest age groups. We show in Fig. 5–B, narrower age groups concerning the youngest patients. This second Figure tells us that the severity related to COVID-19 in children is rather heterogeneous in age, being particularly dangerous for kids below 2 years old (an age segment for which the admissions in intensive care are more common than for patients above 40 years old). Furthermore, these probabilities might be underestimated by the uniform attack rate assumption, since one expects a significantly lower exposure to the virus at these low ages (we will see this confirmed in the data shown in Fig. 6).

**Figure 6:**
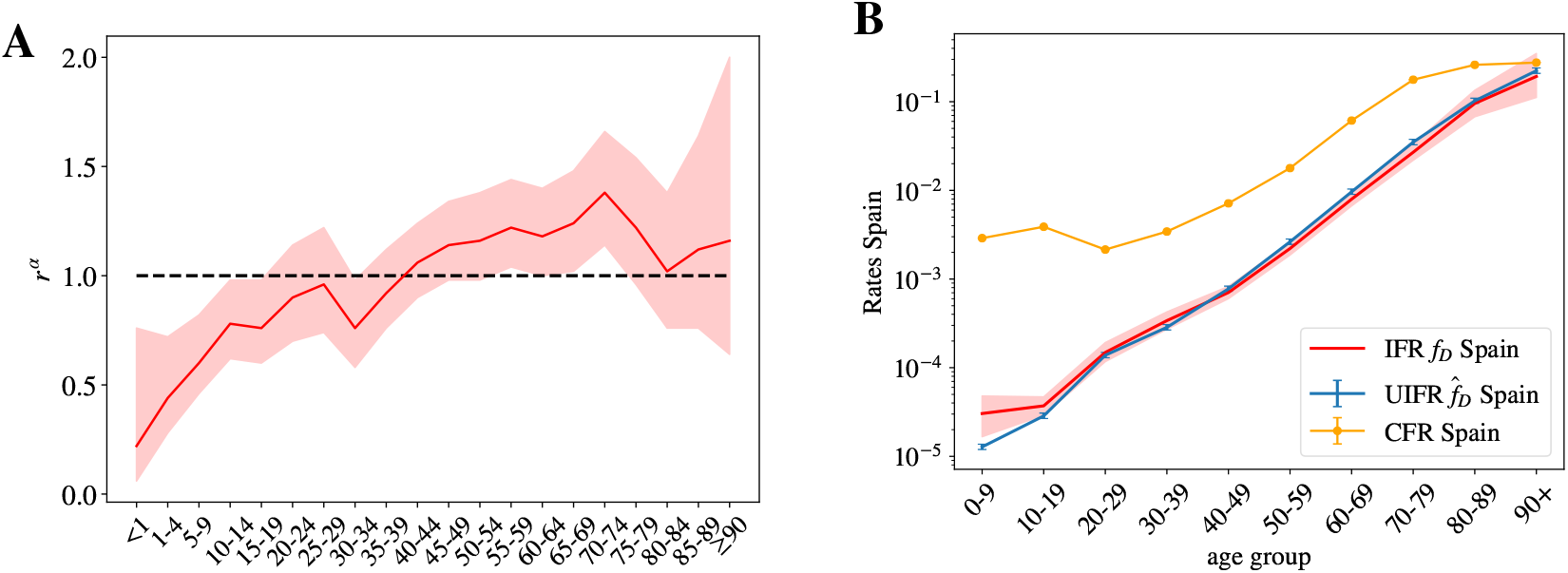
Uniform versus non uniform IFR. A We show the relative risk of infection for each age segment *r_α_* taken from the sero-epidemiological study of the Spanish population [23], using Eq. (3). While the youngest segments of the population seem to be less hit by the virus, the distribution of the infections is rather similar to that of a uniform attack rate, indicated by the dashed line *r_α_* = 1 here. The 95% confidence interval for *r_α_* is indicated by the red shadow. B We show the estimated uniform and nonuniform IFR for Spain and compare it with the case fatality ratio as a function of age. The error for the non-uniform IFR is shown by a red shadow.

## 4 Discussion

### 4.1 On the non-uniform distribution of infections

Our indicator for the fatality ratio 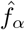 (and the probabilities of presenting different degrees of acuteness) measure how more probable is to die with an age rather than with another in a population, which is not necessarily the true IFR (that is, the probability of dying once infected, our *f_α_* in (2)). The two observables are only equal if the contagion is uniform among all age segments of the population (we recall that, in our definition, 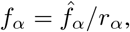 and uniform attack rate implies *r_α_* = 1). In other words, with our approach we are not able to distinguish if the mortality is low in a particular age segment because (i) the disease is mild at these ages (low *f_α_*) or (ii) because this age segment is rarely infected (low exposure, *r_α_ ≪* 1 in Eq. (3)). Previous studies estimating the IFR per age group, for instance Ref. [10], assumed a uniform spread of the virus, something that seems justified by contagion dynamics studies [20].

The sero-epidemiological study [23], gives also some clues about this point, because it also estimates the attack rate for different age groups. We can extract our *r_α_* from this attack rate (we recall that *r_α_I/N*, with *N* the country-population). We show the values we obtain in Fig. 6–A. The measures only report a significantly lower spread among children (which might be related to the closure of the schools during the lock-down), but for the rest of the ages the distribution is not so far away from the uniform attack rate. In any case, no exponentially increasing attack rate with age is found to balance the strong increase of the fatality with age. However, the much lower exposure of the kids to the virus tells us that the probability estimation of Fig. 5–B might be underestimated in that age segment, something that could change the overall picture of the severity of COVID-19 in babies, that might be similar to that of the adults. The change of tendency of the severity with age in the case of infants could related with the suspected connection between the COVID-19 and Kawasaki diseases [32, 33, 34].

We can nevertheless compute the real (non-uniform) IFR using these values for *r_α_* for the Spanish data, and compare it with our previous estimation. We show the results in Fig. 6–B. As shown, both estimations are essentially compatible for all the age segments, which lends confidence to our previous results. The real fatalities will slightly change once the effect of the non-uniform attack rate is included, but we do not expect these non-uniform fatalities to change drastically with respect to the uniform estimations we gave above.

### 4.2 On the under-counting of deaths

As discussed above, we expect that the number of deaths associated to COVID-19 to be underestimated in the official statistics, specially on what concerns to elderly people. Now we try to give an estimate of how much. The collapse of Fig. 3–B shows us that Norway reports a noticeably higher number of deaths in the age segment above 70 years old with respect to other age groups. We believe that their counting is more accurate than for the rest for two reasons. First, because the Norwegian authorities report the deaths (of patients tested positive for COVID-19) occurring everywhere: hospitals (38%), caring and retirement houses (59%) and homes (2%). And second, because the country has been much less affected than the rest (Norway has reported only 235 deaths so far), which means that they are much better equipped to properly detect and treat infections. For this reason, we can use the Norwegian measures give a quantitative estimate to our under-determination of the mortality among the elderly (70–79: 22%, 80–89: 40% and 90+: 86%). We show in Fig. 4–B, that this simple correction allows us to predict correctly the measured prevalence of antibodies within the population in the oldest age segments in the canton of Geneva (Switzerland) [24]

Yet, from this comparison with the Norwegian data we can only argue in terms of the scaling of an age segment with respect to the other, but not on the factor common to all age segments. For this, we can use the comparison between our estimation for the UIFR based on official COVID-19 and the one extracted using the counting of deaths having COVID-19 mentioned in the death certificate. The sero-prevalence study [23] estimated that a 11.3% of the population of the Community of Madrid had been infected, so we can use this number of infected to estimate the fatality of the region. Such a fatality ratio has to be regarded as an upper limit of the real fatality ratio, because “suspicion of COVID-19” probably encompasses many other respiratory diseases. We show this fatality compared to our previous estimation, and the estimation after correcting the under-counting of the oldest segments (using the Norwegian death data) in Fig. S4. We observe that, firstly, the correction introduced for the elderly segments is in perfect agreement with the scaling observed in the Madrid’s data, with attaches confidence to this correction, and second, that Madrid estimation is around 35% larger than our previous estimation for the other ages. This estimation gives us an upper limit of the real mortality, which means that it allows us to estimate the maximum error of the predictions given up to now (since the real mortality must be between that value and this over-estimated one). We show these estimations in Table 2 and Fig. 4–A after taking these effects of under-counting into account.

We can use these corrections to estimate the number of unreported deaths for each of the countries considered and the values of the UIFR per age to compute the global IFR of each country. We show this data in Table 3. Considering that a lower diffusion of the virus among the elderly would result also in a lower apparent mortality in these groups, we give also the expected total IFR if the actual counting were perfect (left side of the parenthesis), and if a constant 35% of under-counting was present in all the age groups (right-side of the parenthesis).

**Table 3:** Country-dependent estimates. We estimate the percentage of unreported number of deaths for each country together with the expected fatality ratio once included these estimated missing deaths. In the parenthesis we include the expected values if the current death counting was perfect (no missing deaths, left side of the parenthesis) and if heavy under-counting was present, such as the one observed when comparing with number of deaths with COVID-19 in the death certificate (right side of the parenthesis).^*∗*^France numbers were computed using only the deaths occurring in hospital facilities, which means that a 58% of under-counting is already confirmed with the counting of deaths occurring in care-houses. We cannot correct the minimum IFR because we do not have the age profile of these deaths.

|  | % of missing deaths | % total IFR |
| --- | --- | --- |
| Spain | 38.%(0-86) | 1.6%(1.1-2.1) |
| Portugal | 9.1%(0-47) | 1.3%(1.2-1.8) |
| Norway | 0%(0-33) | 1.2%(1.2-1.6) |
| Korea | 16.%(0-57) | 0.87%(0.70-1.2) |
| Italy | 61.%(0-120) | 1.8%(0.98-2.4) |
| Germany | 32.%(0-78) | 1.6%(1.1-2.1) |
| France* | 110%(0-190) | 1.6%(0.84-2.2) |
| England | 79.%(0-140) | 1.3%(0.88-1.8) |
| Denmark | 29.%(0-74) | 1.3%(0.97-1.7) |

### 4.3 On the overall infection fatality ratios and demographics

The values of Table 3 shows us that the global fatality of the disease depends strongly on the demographics pyramid of each country, which is the consequence of the nearly exponential dependence of the UIFR with age. In fact, we can use the average values given in Table 2 to explore how the global IFR would change in different parts of the world just because of a different distribution of the number of citizens with age (that is, leaving aside the differences related to the different health systems). While for Italy the IFR is expected to be 1.8%, the same age profile predicts a 0.62% IFR in China (extremely similar to the one estimated in Ref. [10]) or a 0.14% in middle Africa, which could explain, partially, why the outbreaks are significantly less important there than in Europe (where the overall IFR would be 1.38%).

## 5 Conclusions

We have studied the scaling of the accumulated number of deaths related to COVID-19 with age in different countries. After normalizing these numbers by the fraction of people with that age over the entire population, we observe that the lethality of the disease grows (almost) exponentially with age, expanding over almost 5 orders of magnitude between the 0–9 and 90+ age segments. In addition, we show that this scaling with age is essentially country independent for ages under 70 years old. We estimate that the differences observed over this age are mostly related to different levels of under-counting of deaths of elderly people. The collapse of the mortality data allows us establish direct correspondences between the accumulated number of infections occurred in each country since the beginning of the outbreak.

At a second stage, we use the Spanish survey of the sero-prevalence anti-SARS-CoV-2 antibodies in the Spanish population [23] to fix the scale between the number of infections and the number of deaths, which allows us to estimate the COVID-19 infection fatality ratio as function of age (under the assumption of uniform attack rate). We validate these numbers with an analogous prevalence survey of the Genova canton [24]. We also show that, when applied to the COVID-19 death profile of New York City, our predictions are not compatible with the antibody rates estimated by the New York State [35]. This observation suggests that either the real immunity rate is much higher (and reached herd immunity levels) or the fatality ratio has been significantly higher in New York City than in Spain or Geneva, a discrepancy that might be related to a different health system or a collapse of the sanitary system during the worse moments of the epidemics. The scale of the number of infections allows us to compute as well the probability (if infected) of being classified a case, hospitalized, admitted in intensive care units or dying in Spain. The results show a clear growth of all degrees of severity with age, with the notable exception of the infections in patients below 2 years old that lead to much more complications than for older young patients, a situation that could be aggravated by the low exposure of this population to the virus during the lock-down measures.

We further discuss the validity of the uniform attack rate hypothesis using the age distribution of the antibody rates in the Spanish sero-epidemiological study, concluding that even if differences of exposure of the virus between ages are observed, differences do not change qualitatively our estimations for the infection fatality ratio. However, the low attack rate measured among babies warns us that the fatality rate below 2 years old might be importantly underestimated.

We use information concerning the number of death certificates where COVID-19 was referred as possible death cause to show that the under-counting of deaths is a problem that mostly concerns the deaths of old patients. We use the scaling of the mortality with age in Norway to estimate the real fatality ratio of the elderly age segments (in other words, reverse the under-counting). We then validate these estimations with the age profile of deaths in the canton of Geneva and of the deaths certificates in the Community of Madrid.

Finally, our analysis relies exclusively on public statics’ data and can easily be updated as more accurate information is available (for instance regarding the attack rates in different countries or better estimations of the total number of infections). In addition, if consolidated, the probabilities and the approach explained here, can be easily used to estimate the degree of penetration of the SARS-CoV-2 in different cities, regions, or countries, and to track the evolution of the pandemics. Furthermore, we only analyzed the changes of the total mortality with age, but the socio-economical environment of the patients plays also an important role. This study could be generalized to include such variables.

## 6 Methods and Database

### 6.1 Age profile of the COVID-19 deaths

The information about the distribution of the number deaths associated to COVID-19 with age in different countries is taken from the database prepared by the “Institut national d’études démographiques (Ined)” (France) freely available for scientific use at the website https:/dc-covid.site.ined.fr/fr/donnees/. For the rest of epidemic’s measures in Spain (cases, hospitalizations, entries to intensive care unit, we used the COVID-19 datadista database [36]. In both cases, these databases collect together the official information published by each country’s health authorities. The data used corresponds to the 21st and 22nd of May. Some countries do not give the age profile of the total number of deaths, only for a sub-group of the total. If this is the case, we assume a uniform sampling of the ages in all the age segments, and we renormalize all the *D_α_* so that the sum of the deaths over all the age groups matches the total number of deaths published by each country on the 22nd of May of 2020. For the distribution of COVID-19 deaths with age by department in France, we used the data furnished by Santé Publique France, in particular the “donnees-hospitalieres-classe-age” available at the Données hospitalières relatives à l’épidémie de COVID-19 website.

The information about the COVID-19 deaths in the Canton of Geneva is taken from the “N. 5 – 18 au 24 mai 2020” report in the République et canton de Genève website. The information about the deaths in New York city is taken from the “Total Deaths” reports of NYC health website,

The data concerning deaths mentioning COVID-19 in the death certificate was taken from the “up to week ending the 22nd of May” report in the ONS website (England and Wales) and the “Informe de situación 22 de mayo 2020” from Comunidad de Madrid website. The age distribution of the official data (to generate Fig. 1) is taken for (England only) from the Ined database (which is extracted from the daily report of the National Health Service that includes only deaths tested positive for Covid-19 occurred in hospitals only). In order to account for the deaths in Wales, we multiplied the English distribution by 1.05 (Wales deaths represent a 5% of the sum of the deaths of Wales and England in the ONS report). In order to estimate the official age distribution of deaths in Madrid, we renormalized the national age distribution of accumulated deaths by the official accumulated number of Madrid at the 14th and 22nd of May. This is a reasonable approximation considering that almost a third of the total COVID-19 deaths in Spain occurred in Madrid.

### 6.2 Demographics information

For the demographics distribution, we used the data available at the Ined database which corresponds to the last distribution published by each country official statistics’ agencies, and the database from the ‘World Population Prospects’ of the United Nations https://population.un.org/wpp/Download/Standard/Population/ for the discussions about demography distribution in other parts of the world. The demographics of the canton of Geneva were extracted from Statistiques cantonales in the République et canton de Genève website. For the demographics of New York City we used the data published in the NYCdata website from 2016.

## Data Availability

All the data used is public and freely available at the websites referenced in the paper

## 7 Acknowledgments

I would like to thank Aurélien Decelle, Luca Leuzzi, Enzo Marinari, Giorgio Parisi, Federico Ricci-Tersenghi, Riccardo Spezia and Francesco Zamponi for useful and interesting discussions, and to Elisabeth Agoritsas, Ada Altieri and Marco Baity-Jesi and David Yllanes for a critical and constructive read of the manuscript.

I also thank the Ministerio de Economía, Industria y Competitividad (MINECO) (Spain) through Grant PGC2018–094684-B-C21 (also partly funded by the EU through the FEDER program), for partial financial support.

---

**Figure S1:**
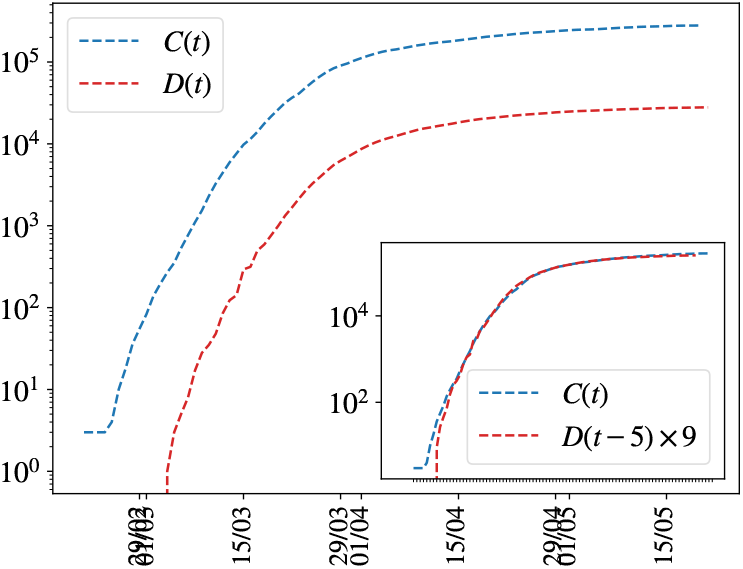
Simple scaling relation linking the evolution of the accumulated number of deaths and the cases with time. We show the evolution with time of the accumulated total number of official COVID-19 cases and deaths in Spain. In the inset the deaths’ curve is displayed 5 days backwards in time and multiplied by 9, following very precisely the cases’ evolution once it surpassed approximately the 100 cases.

**Figure S2:**
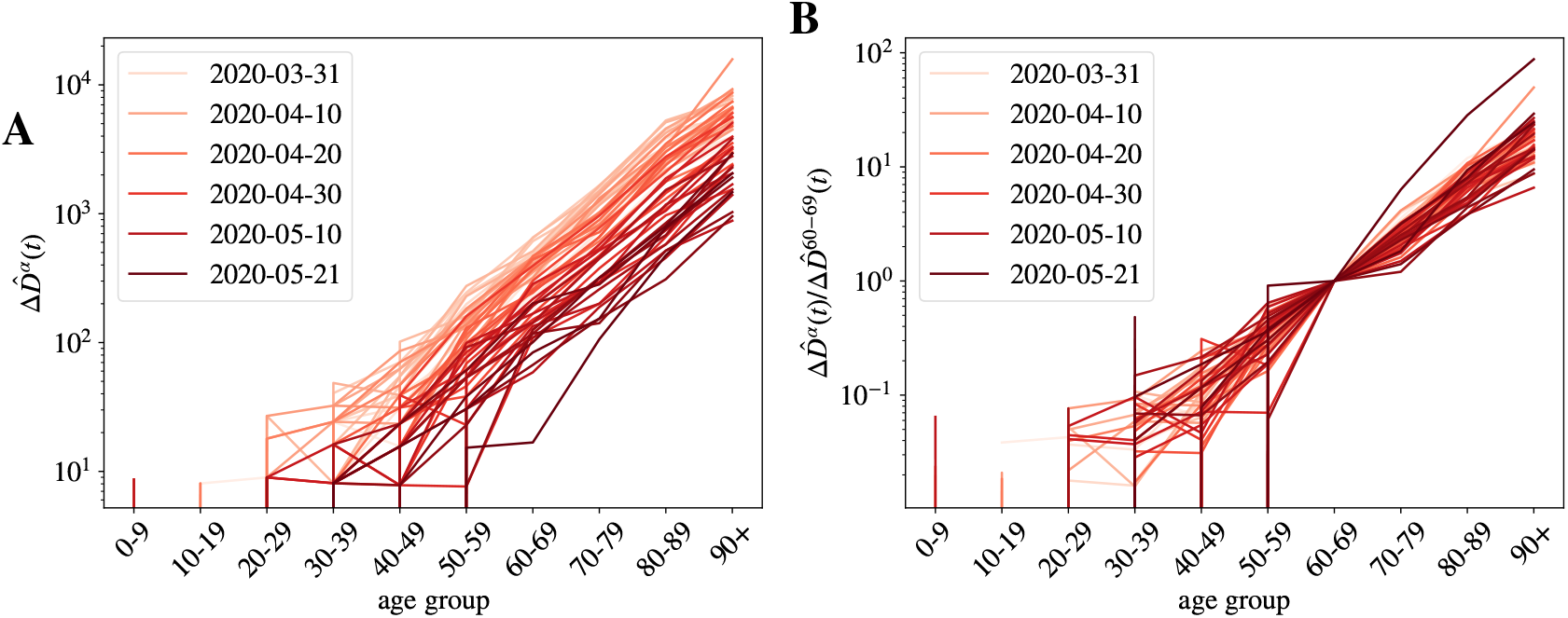
Daily normalized number of deaths registered in French hospitals as function of age and time. A We show the daily measures of deaths for age group *α* (normalized by the population density at this group), Δ ^*D_α_*(*t*), for different dates. The darker the color, the more recent the measure. In B we show the collapse of the data when we normalize the data with the numbers of group 60–69 years old. Distinct date data collapse worse in a single curve than in the case of the accumulated number of deaths in Fig. 2 because being the daily measures smaller, the fluctuations are much larger, yest, we do not observe any systematic change of the attack risk *r_α_* with time.

**Figure S3:**
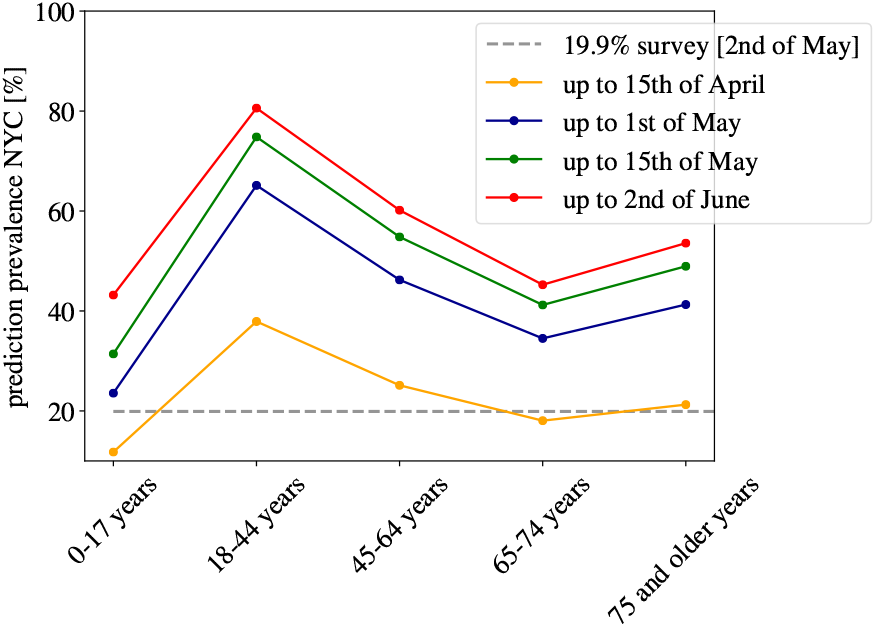
Predictions for the sero-prevalence in New York City. We show our predictions for the sero-prevalence presence in New York City using the death age profile published at different dates and the IFR of Table 2 (without under-counting corrections). Our predictions are significantly higher than the results of the sero-epidemiological survey announced by the New York State Governor the 2nd of May of 2020.

**Figure S4:**
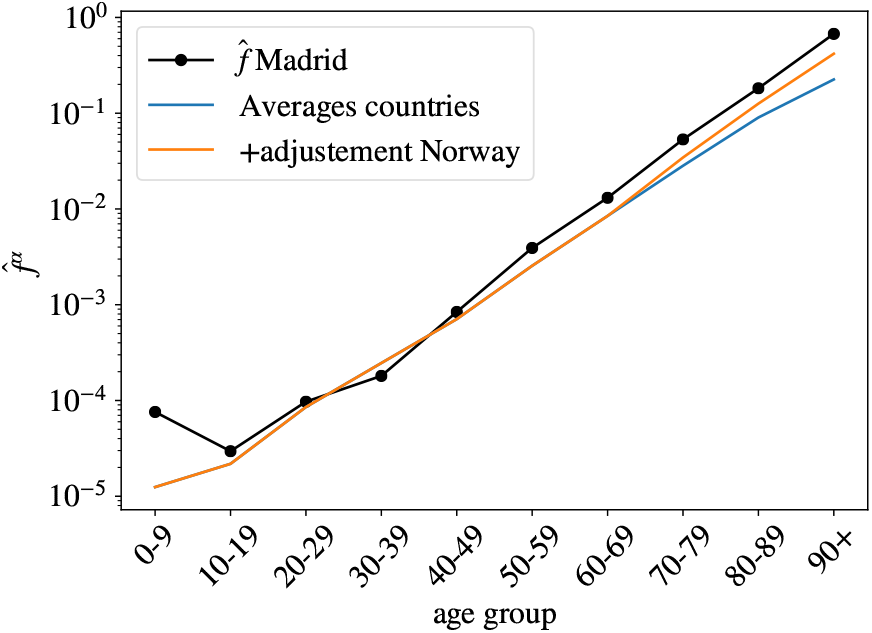
Estimation of the uniform infection fatality rate by age for the Community of Madrid using the number of deaths where COVID-19 was mentioned in the death certificate (black dots), compared with our estimation of the UIFR extracted from the average of several countries (blue line) and the same estimation where the fatality of the oldest segments was adjusted to take into account the systematic under-counting of elderly deaths (estimated using the Norwegian distribution of deaths with age). We see that this correction match very well the scaling observed in Madrid’s data.

## Footnotes

1 In general, Δ depends on *α*, but we omit it here for simplicity because the differences are extremely subtle at this time of the outbreak.

